# Exploring the application of deep learning methods for polygenic risk score estimation

**DOI:** 10.1101/2023.12.14.23299972

**Authors:** Steven Squires, Michael N. Weedon, Richard A. Oram

## Abstract

**Background:** Polygenic risk scores (PRS) summarise genetic information into a single number with multiple clinical and research uses. Machine learning (ML) has revolutionised a diverse set of fields, however, the impact of ML on genomics in general, and PRSs in particular, has been less significant. We explore how ML can improve the generation of PRSs.

**Methods:** We train ML models on known PRSs using UK Biobank data. We explore whether the models can recreate human programmed PRSs, including using a single model to generate multiple PRSs, and the difficulty in using ML for PRS generation. We also investigate how ML can compensate for missing data and the constraints on performance.

**Results:** We demonstrate almost perfect generation of PRSs, including when using one model to predict multiple scores, and with little loss of performance with reduced quantity of training data. For an example set of missing SNPs the MLP produces predictions that enable separation of cases from population samples with an area under the receiver operating characteristic curve of 0.847 (95% CI: 0.828-0.864) compared to 0.798 (95% CI: 0.779-0.818) for the PRS. We provide evidence that input information is the limiting factor of further improvement.

**Conclusions:** ML can accurately generate PRSs, including with one model for multiple PRSs. The models are transferable and have high longevity. With certain missing SNPs the ML models can statistically significantly improve on normal PRS generation. Models trained are probably at the edge of performance and further improvements likely require use of additional input data.

Code is available at https://github.com/stevensquires/

## 1) Introduction

Polygenic risk scores (PRSs), also known as genetic risk scores, are a method of summarising risk information from the genome into a single number (1). PRSs show high discrimination for certain diseases and can be used for various clinical and research purposes including stratifying people into risk classes (2), enabling improved targeting of diagnostic methods and predicting disease progression (3).

A PRS is typically produced from a genome wide association study (GWAS) (4) aimed at assessing which single nucleotide polymorphisms (SNPs) are associated with a partition of the samples. The GWAS provides the statistical significance of the SNPs along with an associated weight which can be used in the PRS. A common approach to produce a PRS is to take the statistically significant SNPs with the associated weights and sum the product of the weights with the effect allele count. Alternative methods can utilise more complex functions such as the inclusion of pairwise interaction terms (5).

Machine learning has been successfully applied to medical and biological domains. Examples include the use of deep learning in medical imaging (6), the prediction of progression of disease (7), and the discovery of protein structure (8). However, these approaches have not worked as well in genetics and genomics in part due to the tabular nature of genetic data (9) alongside the challenges of high dimensionality. There is potential for machine learning to have a substantial impact on genomics, as it is having elsewhere, if appropriate methodologies can be found.

In this paper we explore how machine learning can be used in the generation of PRSs. We consider three overall aims: 1) to investigate the use of deep learning to replicate human programmed PRSs; 2) to explore whether deep learning can compensate for missing input SNPs; 3) to examine the limitations of the deep learning models to compensate for missing SNPs. To test whether deep learning can generate PRSs we take four PRSs, all with non-linearities due to interaction terms, and train deep models on the labelled outputs using a large dataset.

There are several potential advantages of using deep learning to generate PRSs. One is the transferability of the models to researchers and clinicians. Complex PRSs (with non-linearities) are typically generated by specific code written in a particular programming language. With deep learning models the predictions are sets of matrix multiplications with trained weights which can be stored in formats accessible to people using many analysis platforms or programming languages. This approach also provides longevity as the matrices can be stored in formats which will still be readable as programming languages change with time. Furthermore, if we can train a single model to accurately predict multiple PRSs it simplifies the generation of these complex PRSs.

Another potential use of deep learning based PRSs is that they can compress the data into latent representations that may be of use to further prediction models. A PRS is a scalar, and it may be that a vector-based compression may allow for improved predictions by models which combine multiple factors into a risk model.

The training of a deep network on PRSs may also function as an effective self-supervised learning (SSL) pretext task. SSL is becoming one of the most important approaches in deep learning (10) and a crucial part of the process is the choice of pretext tasks. By training models on the PRSs this may allow the model to find a set of parameters which can then allow improved training on the specific desired task. The use of SSL has already been demonstrated in genomics (11) and many further uses would be expected.

The second aim is to be able to generate a complex PRS from subsets of the input SNPs using machine learning approaches. The values of SNPs can be missing, or of poor quality, and this will reduce the performance of the PRS. We consider three potential mechanisms a model could use to accurately estimate the PRS without all the input SNPs. The first is that reweighting the remaining SNPs may compensate for the missing SNPs. The second is that the values of the missing SNPs can be estimated. The third is that there are patterns in values of the remaining SNPs that can be leveraged to predict what the final PRS should be.

We explore three models in this study. The first is a linear predictor which tests the extent to which the reweighting of the SNPs can enable the original PRS to be reproduced. The second method is to use a variant on an autoencoder (12) which can take in the remaining SNPs and outputs the prediction of the values of the original SNPs. The third is a multi-layer perceptron (MLP) (13) which should cover all three hypotheses and is the main model we focus on throughout the paper.

The third aim of this study is to probe the limitations of deep learning to compensate for missing SNPs. We take a SNP subset and train multiple models with different architectures, training approaches and encodings of the inputs. If the constraints on performance are due to the quality of the models we should expect to observe improvements in performance over a base model. Alternatively, if the limitation is due to lack of information in the input data there should be little improvement when exploring different model variants.

## 2) Methodology

### 2.1 Polygenic risk scores (PRSs)

We consider four PRSs for two diseases. Three are for risk score estimation for type 1 diabetes (T1D) and the fourth is for Celiac disease. We primarily study a T1D PRS generated from 67 input SNPs (5) which we label as T1D67. This PRS includes both a standard additive component alongside a more complicated set of interaction terms which describes important relationships within the human leukocyte antigen (HLA) region of chromosome 6.

The other two T1D PRSs use ten SNPs, denoted T1D10, and thirty SNPs, denoted T1D30 (14), respectively. Both include an additive part alongside an interaction term based on two SNPs. The fourth PRS is produced with 42 SNPs for Celiac disease, denoted CD42, and contains similar interaction terms to the T1D67 PRS.

### 2.2) Data and pre-processing

For training and testing we utilise the UK Biobank (UKBB) genomic data imputed using Minimac servers (15) with a combination of the 1000G panel data (16) and the Haplotype Reference Consortium panels (17). The dataset consists of 487,409 samples which we partition in slightly different ways to include the T1D and Celiac samples as a separate test set to check the discrimination capacity of the models. We use 387 T1D and 1,184 Celiac cases.

When building models individually on the T1D PRSs (T1D10, T1D30 and T1D67) we partition the 387 T1D cases into a separate test set. The rest of the samples are randomly assigned into 70% training (340,915), 15% validation (73,053) and 15% testing (73,054) sets. When building models to predict the CD42 PRS we partition the 1,184 Celiac cases into a test set then randomly split the rest of the data into 70% training (340,357), 15% validation (72,933) and 15% testing set (72,935).

We also produce a single model which can predict all four PRSs, for which we hold out both the 1,184 Celiac disease and 387 T1D cases in two separate test sets. There are 8 samples in both Celiac and T1D test sets. The remaining data is split into 70% training (340,092), 15% validation (72,876) and a second testing set (72,878). For the two test sets we denote the samples with T1D or Celiac as case test sets and the other as population test sets.

For the input data we convert all SNP values to be based on the effect allele, so that a 0 represents no effect allele being present. Any missing values were set to 0 before training and testing of our models. The four PRSs were generated for all 487,409 samples. These scores are our ground truth label for training of the models and assessment of their quality.

In addition to the UKBB data we use data from the Type 1 Diabetes Genetics Consortium (T1DGC) study (18) which consists of 9,413 non-T1D controls and 6,666 T1D cases. We use this dataset for inference and perform no training or model selection on it.

### 2.3 Generation of PRSs from neural networks

Our main approach is to use an MLP for generation of PRSs. A representative (not showing actual number of layers or neurons) diagram of this type of model is shown in Figure 1 with five neurons in the input layer (*IL*), two hidden layers (denoted by *HL*_*1*_ and *HL*_*2*_) with 4 and 3 neurons respectively and a single neuron in the output layer. The model is fully connected with all weights learnable during training. The basic MLP consists of three hidden layers each with 50, 30 and 20 neurons respectively. The input number of neurons is defined as the number of SNPs in the PRS (or the number of subset SNPs being investigated). We use rectified linear units (ReLUs) (19) as the activation function for the hidden layer neurons and no additional activation function for the final output neuron. We use the mean squared error (MSE) objective function 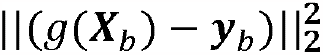 to train the model where we denote the batch by subscript b and g is the function produced by the MLP. We refer to this model as the standard MLP.

**Figure 1.**
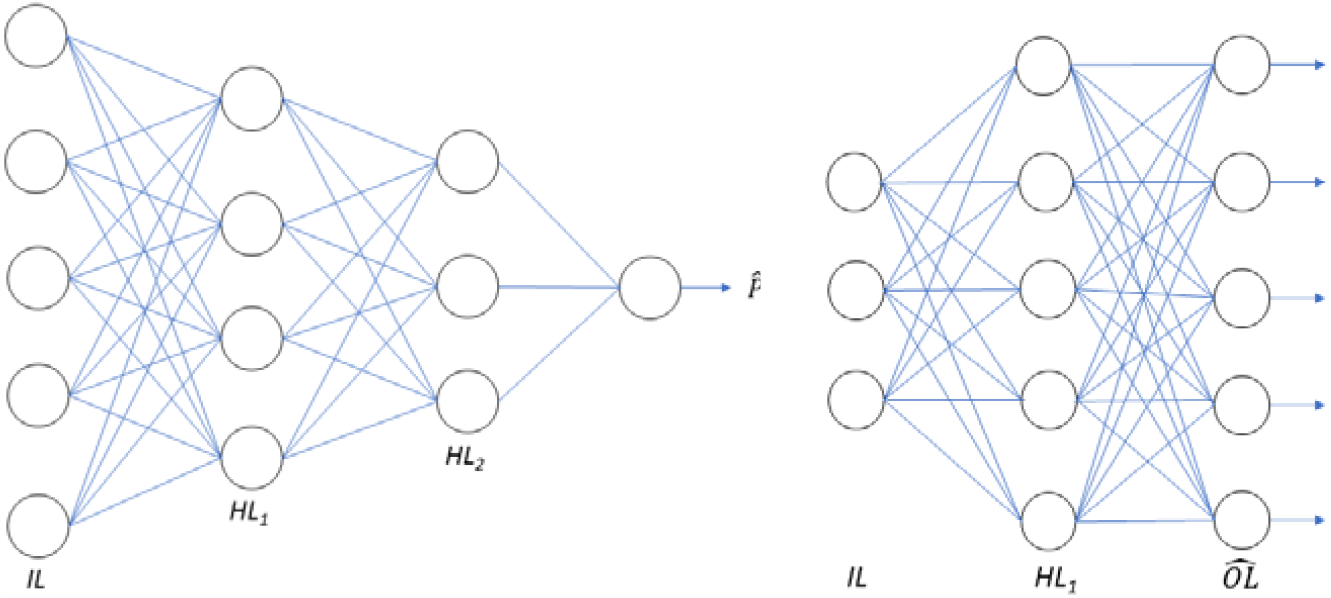
Schematic diagrams of the structure of the MLP (left) and AE (right). Each circle represents one neuron and the lines represent the links between the neurons on adjacent layers. IL stands for “input layer” where each neurons represents an input SNP, HL is the “hidden layer”. For the MLP (left) the final output is the prediction of the PRS 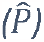 while for the AE (right) the output layer 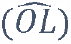 is the prediction of the original 67 input SNPs.

A potential use case for the neural network approach is to provide one model which outputs multiple PRSs. This is demonstrated in Figure 2 where the previous MLP model (in Figure 1) is altered to have four output neurons, each representing one of the four PRSs. The number of inputs is now the combined SNPs required for all four PRSs but otherwise no changes are made from the previous MLP. For clarity we refer to this model as the multi-predictor MLP

**Figure 2.**
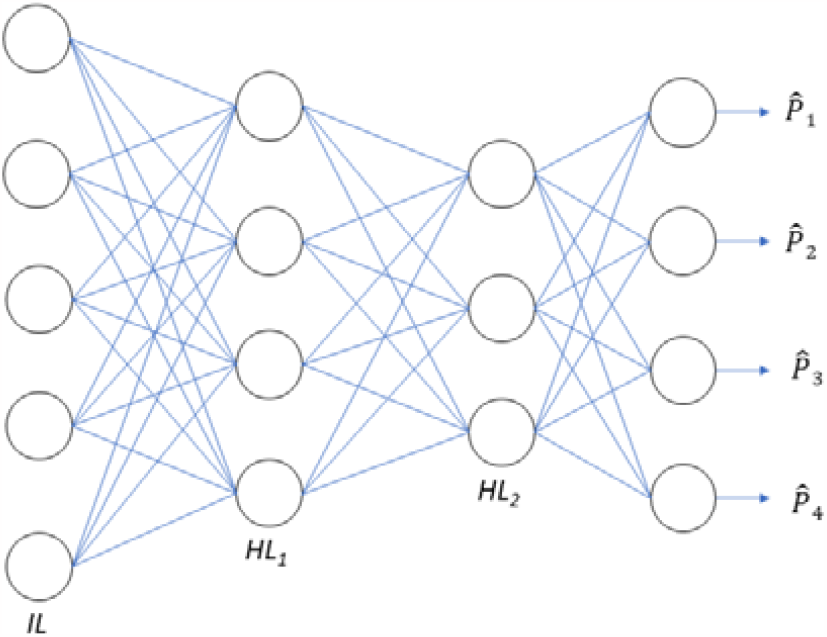
The multi-predictor MLP model schematic. The model design is the same as the standard MLP (in Figure 1). The changes are in the input layer (IL) where the number of SNPs is increased to include all necessary SNPs for all the desired models and in the output layer where each output neuron represents one of the PRSs (in this case with four).

To act as a model comparison to the MLP, and to test the reweighting hypothesis when we examine reduced number of input SNPs, we train a linear predictor, using Sci-Kit Learn (20). The number of samples is much larger than the number of features but we also applied an elastic net form of regularisation (21) to the trained models to check if performance improved. The linear predictor is given by 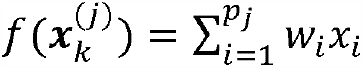, where *p*_*j*_ is the number of SNPs for the j^th^ set of SNPs, 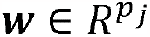is the vector of weights to be found and 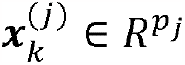 is the input SNP values for the k^th^ sample. The elastic net is trained to minimise 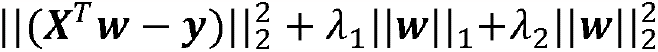 where the training samples (with number *n*_*tr*_) have been stacked into matrix 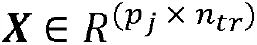. A grid search over *λ*_l_ and *λ*_2_ was performed and the MSE assessed on the validation data. Regularisation added no improved performance on the validation set and we reverted to using a direct linear predictor with no regularisation, therefore optimising 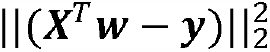.

We also developed a variant of an autoencoder (AE) to predict missing SNP values when we reduce the number of input SNPs. A representation of this model is shown in Figure 1 with an example three neurons in the input layer (*IL*), 5 neurons in the hidden layer (*HL*) and 5 neurons in the output layer 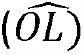. The actual AE model is designed with two hidden layers each with 67 neurons connected to an output layer with 67 neurons each one representing one of the original SNPs. The number of neurons in the input layer is defined by the number of input SNPs. The activation function of the neurons in the hidden layers is a ReLU and in the final layer a sigmoid multiplied by two to allow prediction of the SNP value (0, 1, 2).

The AE is trained to produce a 67-neuron output with the MSE objective function 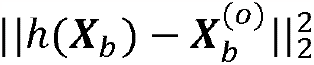, where h() is the function produced by the AE, ***X***_*b*_ is the stacked batch of inputs where each ***x*** is the *p*_*i*_ dimensional vector for that number of SNPs and 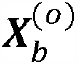 is the batch of original inputs with all the SNPs intact. The AE output is the estimate of the original SNP values. To generate the PRS estimate for each input 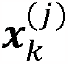 we apply the p_0_ -SNP (all 67 SNPs) trained MLP, with function g_0_, to the output of the AE to give our estimate of the PRS: 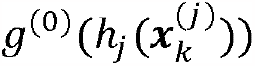. We denote the AE followed by MLP method as AE-MLP.

In Figure 3 we show the three functions (MLP, linear predictor and autoencoder followed by MLP) we use for predicting a PRS including when there are missing input SNPs. The subscripts denote the sample and the superscript a set of remaining SNPs with (0) showing all original SNPs.

**Figure 3.**
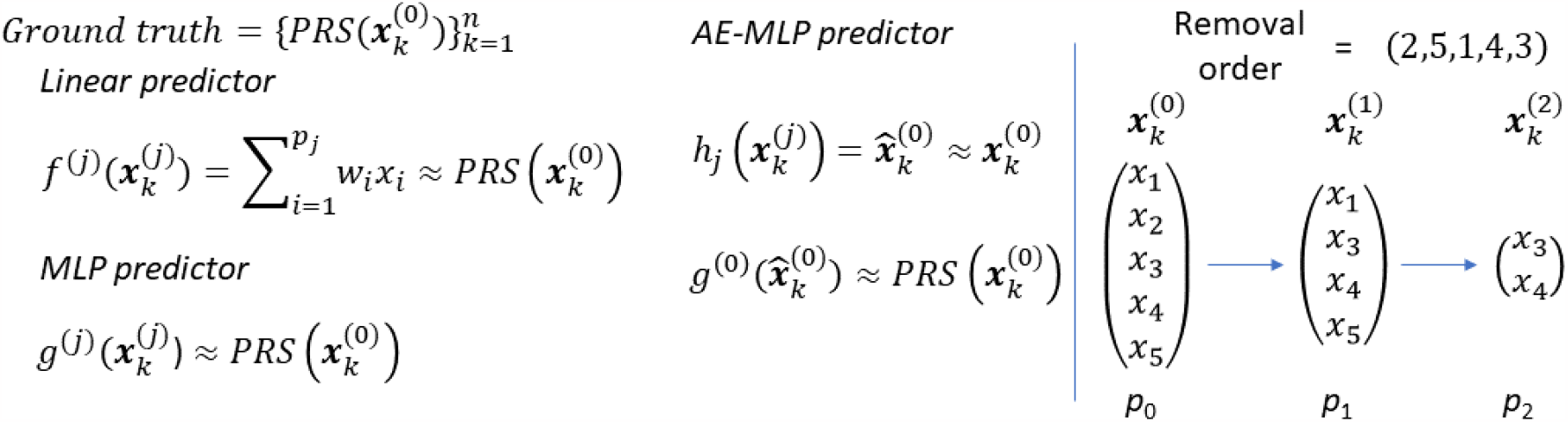
Left) The functions of the three predictors (linear, MLP, AE-MLP) with the functions represented by f(), g() and h(), respectively. Right) The procedure for removing the input SNPs. The removal order is shown at the top and then three versions of the inputs are displayed with the first showing the inclusion of all the SNPs and then the next two with the removed SNPs in order. The subscript k represents the k^th^ sample and the superscript a representation of which SNPs are included/excluded.

We use PyTorch (22) for both the MLP and AE using the Adam (23) optimiser with hyperparameters other than the learning rate left as defaults. The MLP is trained with a learning rate of 10 × 10^−4^ and a batch size of 5 while the AE is trained with a learning rate of 5 × 10^−5^ and a batch size of 5. The batch-sizes and learning rates were selected by assessment of the training and validation plots due to the number of models to be trained they are kept the same for all the basic models.

### 2.4) Estimating the PRS with missing SNPs

One of our objectives is to estimate a PRS without having access to all the required input SNPs. In Figure 3 we show a graphical representation of the procedure. In this toy example there are three subsets with no SNPs removed, one SNP removed, and three SNPs removed, respectively. We show the effect of different removal permutations in the results.

For all choices of removed SNPs we directly generate the PRS without the relevant SNPs to show how the gap between the PRS on all 67 SNPs and the reduced number of SNPs varies. We then train the three models (linear predictor, AE-MLP and MLP) with the reduced number of SNPs as the input and the PRS (generated with the full set of SNPs) as the label.

### 2.5) Additional MLP models for finding model performance limitations

A further objective is to understand whether limitations in the MLP predictive power with missing SNPs is due to the quality of the models or the information content of the inputs. The UKBB dataset is large and the number of inputs small so neither data quantity nor overfitting should limit the model performance. Therefore, if the models cannot recreate the original PRS it should be due either to the model or the amount of information in the input SNPs.

We test whether the constraint on performance is due to the models by exploring variants on the MLP. A MLP is known to be a universal function approximator (24) so can, in principle, find the optimal mapping from a set of inputs to desired output. If we can find no improvement in model performance by searching through different model variant it provides evidence that the constraint is the information content of the input SNPs.

We focus on one specific set of SNPs so that we do not have an unfeasibly large number of models to train. We take the 52 SNPs from the second random permutation (the middle plot of Figure 8) and train multiple model variants altering important aspects of the model to attempt to improve performance.

The choice of hyperparameters can substantially improve model performance, two important ones are batch size and learning rates. Due to the large number of models we needed to train we did not do a search through these and chose a learning rate and batch size which produced training and validation curves with iterations that converge. For this one SNP subset we do a grid search through the learning rates and batch sizes to ascertain whether any improvements can be made. We select the best performing model by its performance on the validation set. In the results these models are referred to as *Hyperparams*.

There may be an issue with model capacity if there are a larger number of required functions needed than is possible with the current model architecture and design. There may also be difficulties with correctly assigning weights throughout the network. To test these issues we create four model architectures by altering the number of layers and number of neurons within each layer leaving the ReLU activation functions in the hidden layers and the linear output neuron the same. The first is the original MLP architecture described earlier with [50, 30, 20] neurons in each hidden layer; the second has one hidden layer with 200 neurons; for the third the number of hidden layers is 7 with [55, 50, 40, 30, 25, 15, 5] neurons within each layer; the final model also has 7 hidden layers with [200, 160, 120, 90, 70, 50, 10] neurons in each layer. Between these different architectures we should be able to assess whether the model capacity or the difficulty in training deeper models produces limitations on predictive quality. In the results section these four model architectures are labelled as *Capacity* 1-4 respectively.

An important advance in improving deep learning models was the use of connections between non-adjacent layers (25). To test if these skip connections can improve performance we add links from the input layer to the final hidden layer (with 20 neurons), keeping the rest of the number of layers and neurons the same as the standard MLP. A schematic of this skip-connection approach is shown in the left plot of Figure 4. These skip connections alter the number of neurons in the last hidden layer from 20 to 20+52. This should ensure that any information lost is returned before the final mapping. In the results this model is labelled *Skip*.

**Figure 4.**
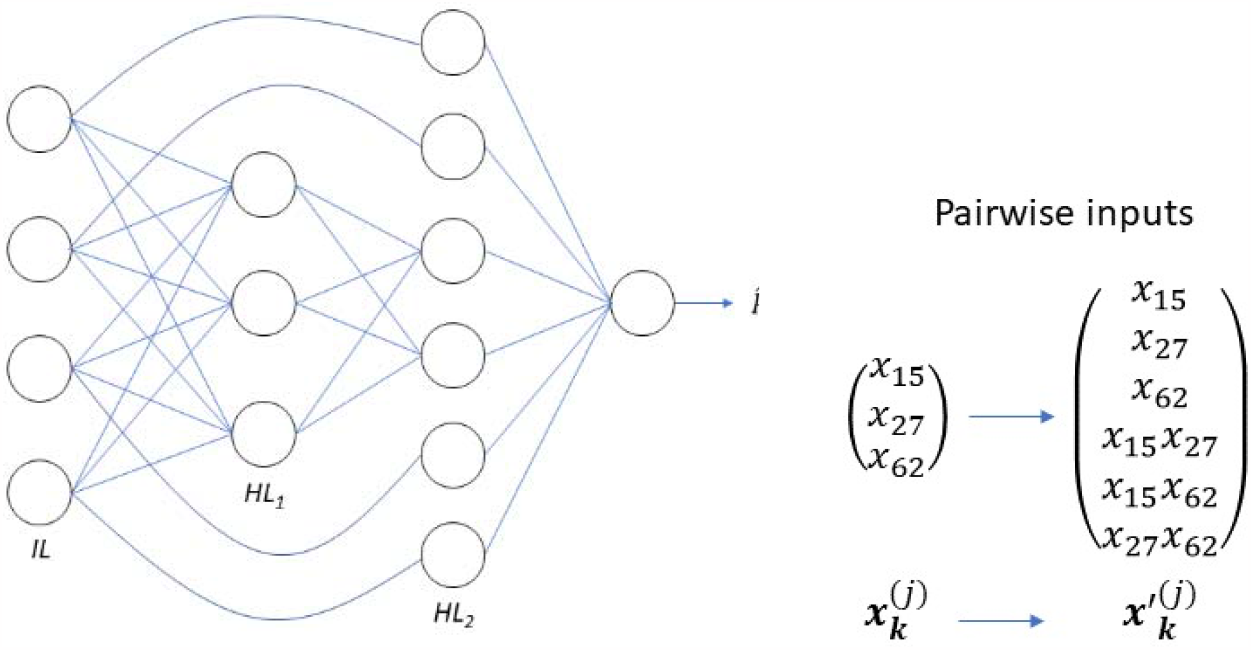
Left) Schematic example of the skip connection model. The input layer is connected to the first hidden layer (HL_1_) as standard but also the input neurons are directly connected to the final hidden layer (HL_2_). There are no weights applied to the skip connections. Right) Example of the expansion of the inputs when including direct pairwise terms. Each input is multiplied individually by all other inputs and then each of those values are concatenated together with the individual values. In the example there are three individual input SNPs which when combined with the pairwise terms creates a 6-dimensional vector.

The final model variant is to alter the input SNPs to include all possible pairwise combinations. We leave in the 52 input SNPs and append all combinations of the pairs multiplied together. This adds in an addition 1,326 input neurons to give a total of 1,378 with the input now altered to (x_1,_x_2_,…x_52_,x_1_x_2,_x_1_x_3_,…,x_51_x_52_). While this approach adds no additional information for the model it should make it easier for it to find relationships between pairwise interactions. The approach is shown in the right of Figure 4. In the results this model is called *Pairwise*.

One of the most effective ways to improve model performance is to utilise ensembles or averages of multiple models, ideally as diverse models as possible. We therefore also explore whether combining the predictions of the different models allows for any improvement in overall performance.

### 2.6) Transferability and longevity of the models

A potential advantage of the neural network approach to PRS generation is the ease of transferring models and their longevity. The importance of these two factors become more significant when the models become more complicated as code complexity will also increase. As PRSs are developed in one coding language they may not be accessible to people not familiar with that specific language. In addition, coding languages and other technology change over time and the code needs to be maintained to ensure it continues to work effectively.

Our MLP is a combination of matrix multiplications with the ReLU function. The standard MLP consists of four matrices with associated bias vectors. To generate the final MLP-PRS 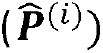 from the input SNP vector from the *i*^*th*^ sample 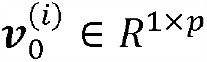 requires four steps:

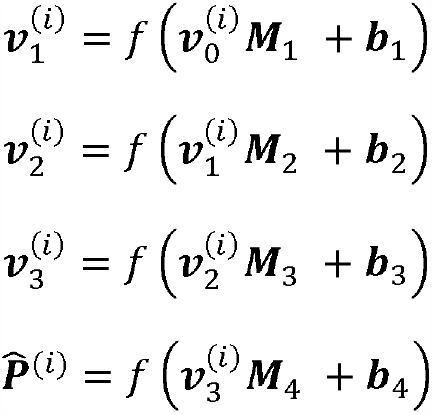

where *f(x)* is the ReLU function *f* (*x*) *= max*(0,*x*)and the matrices are of sizes: 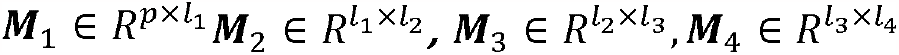 and biases: 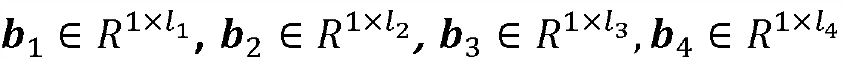 The number of neurons in each matrix is *l*_1_ = 50, *l*_2_ = 30, *l*_3_ = 20 and *l*_4_ = 1.

The storage of the parameters can be done in simple formats such as basic text files which is unlikely to become obsolete so there should be longevity in use of these models. Matrix multiplications are also simple to perform in almost all programming languages and other tools such as spreadsheets so should be highly accessible. Any changes in programming languages can easily be adapted to perform the matrix calculations. The only other information that needs to be sent with the matrices and biases is the list of ordered SNPs to match the input vectors along with the alleles.

## 3) Results

### 3.1 Generation of PRSs using MLPs

In Table 1 we show test set Spearman rank correlation coefficient (*Spearman rank*) and root mean squared errors (RMSE) for the standard MLP for the four PRSs when models are trained separately. For context of the RMSE values the T1D10, T1D30, T1D67 and CD42 mean scores are 9.2, 12.6, 10.1 and 2.4 respectively. We also show the area under the receiver operating characteristic curve (AUC) between the cases and population test sets for the PRS (*PRS AUC*) and the MLP (*MLP AUC*). The MLP can estimate the PRS to a high degree for all the PRSs and produce the same level of discrimination performance.

**Table 1.** Metrics for the four individual MLPs trained separately on the PRSs. We show the Spearman rank correlation coefficient (Spearman Rank) and RMSE between the MLP estimates and PRSs. Also displayed are the area under the receiver operating characteristic curves (AUC) between cases and population test sets for the PRS (PRS AUC) and the MLP (MLP AUC). Confidence intervals are generated by bootstrapping and reported at the 95% level.

| PRS | Spearman rank | RMSE | PRS AUC | MLP AUC |
| --- | --- | --- | --- | --- |
| T1D10 | 0.999 (0.999-0.999) | 0.052 (0.051-0.053) | 0.855 (0.837-0.873) | 0.855 (0.837-0.879) |
| **T1D30** | 1.0 (1.0-1.0) | 0.055 (0.054-0.057) | 0.865 (0.846-0.882) | 0.865 (0.844-0.885) |
| **T1D67** | 0.997 (0.997-0.997) | 0.175 (0.172-0.179) | 0.919 (0.904-0.932) | 0.919 (0.902-0.931) |
| **CD42** | 0.998 (0.998-0.998) | 0.105 (0.102-0.107) | 0.874 (0.866-0.884) | 0.873 (0.861-0.882) |

To explore the robustness of model training we show, in Figure 5, how the validation error falls when training on the T1D67. We show plots of the validation losses against iteration for different learning rates (left), different batchsizes (middle) and different model architectures (right). For both varying learning rates and varying batchsizes the model approaches a similar level of performance. The number of neurons in each layer for the five models is: model 1 (the standard MLP) has (67, 50, 30, 20, 1), model 2 has the same number of layers but fewer neurons (67, 20, 10, 5, 1), model 3 has the same number of layers with more neurons (67, 100, 60, 40, 1), model 4 has only one hidden layer (67, 50, 1), and model 5 has more hidden layers (67, 50, 40, 30, 20, 10, 1). Model four does not find a local minimum comparable to the other models.

**Figure 5.**
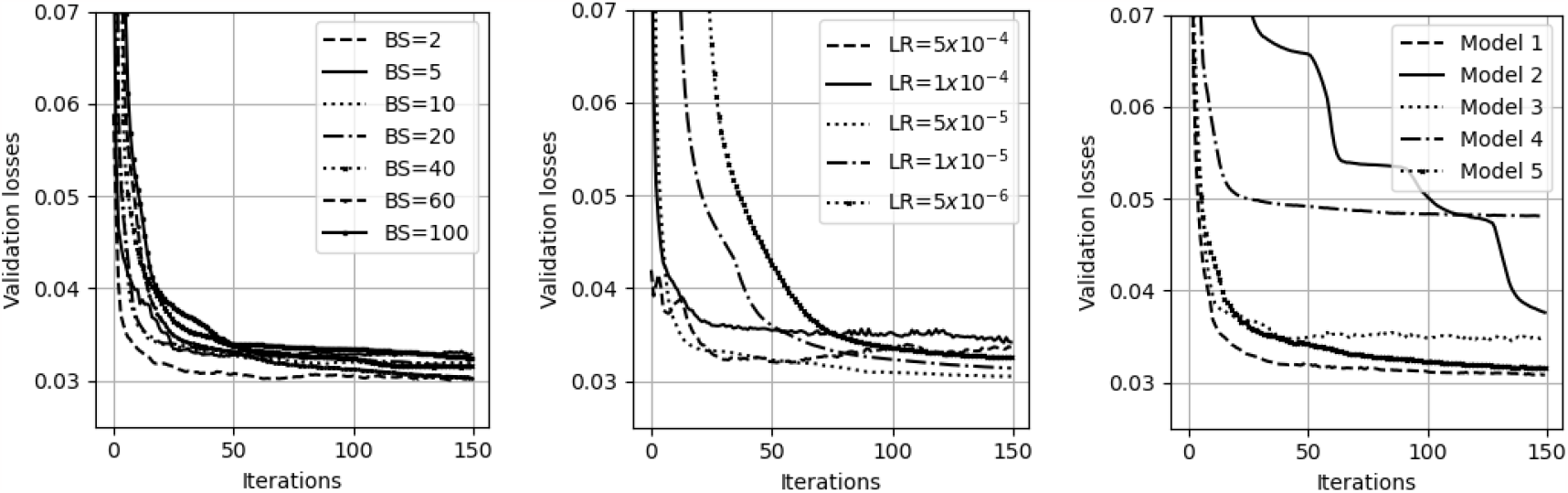
Examples of validation errors in training runs for different hyperparameter choices. Left) Validation losses for different batchsizes (BS) for the standard MLP model, all with a learning rate of Middle) Validation losses for different learning rates (LR) for the standard MLP model with a batchsize of 20. Right) Validation losses for different model architectures all trained with a batchsize of 20 and learning rate of

In Figure 6 we show model performance at estimating the T1D67 with changing size of the training set. The left plot shows the AUC between the cases and population test sets. The middle and right plots show the Spearman rank correlation coefficient and RMSE between the model predictions and the T1D67 on the population test set, respectively. The original dataset has around 400,000 samples but prediction quality is only modestly reduced as the number of samples falls below 10,000.

**Figure 6.**
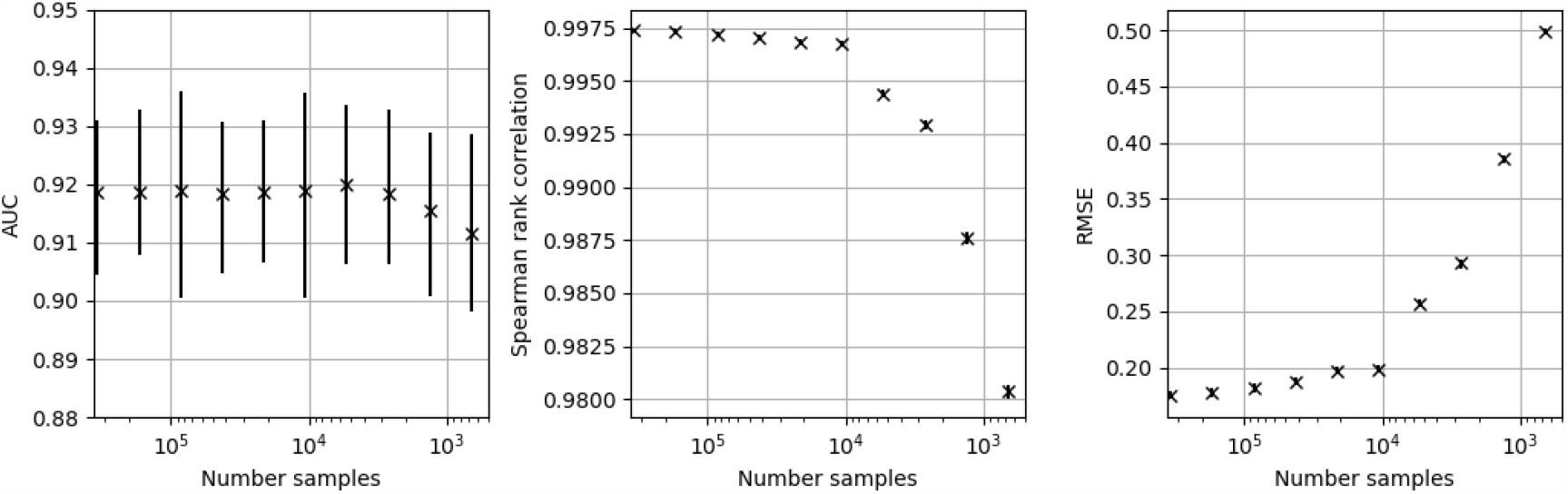
The performance of the standard MLP model at predicting the PRS67 with reduced numbers of training samples. Uncertainty estimates are produced by bootstrapping and reported at the 95% level. Left) The AUC for separation of the test set drawn from the population compared to the 387 T1D samples. Middle) The Spearman rank correlation coefficient for the MLP predictions compared to the PRS67 values for the population test set. Right) The root mean squared error (RMSE) for the MLP predictions compared to the PRS67 values for the population test set.

In Figure 7 we show how one model (the multi-predictor MLP) can estimate all four PRSs. The left plot shows the four validation losses (one for each PRS) during training of the model. The right plot shows the AUCs for the four models compared with the AUCs from the PRS. The prediction performance (as measured by AUC) is the same for the multi-predictor MLP as the four individual PRSs.

**Figure 7.**
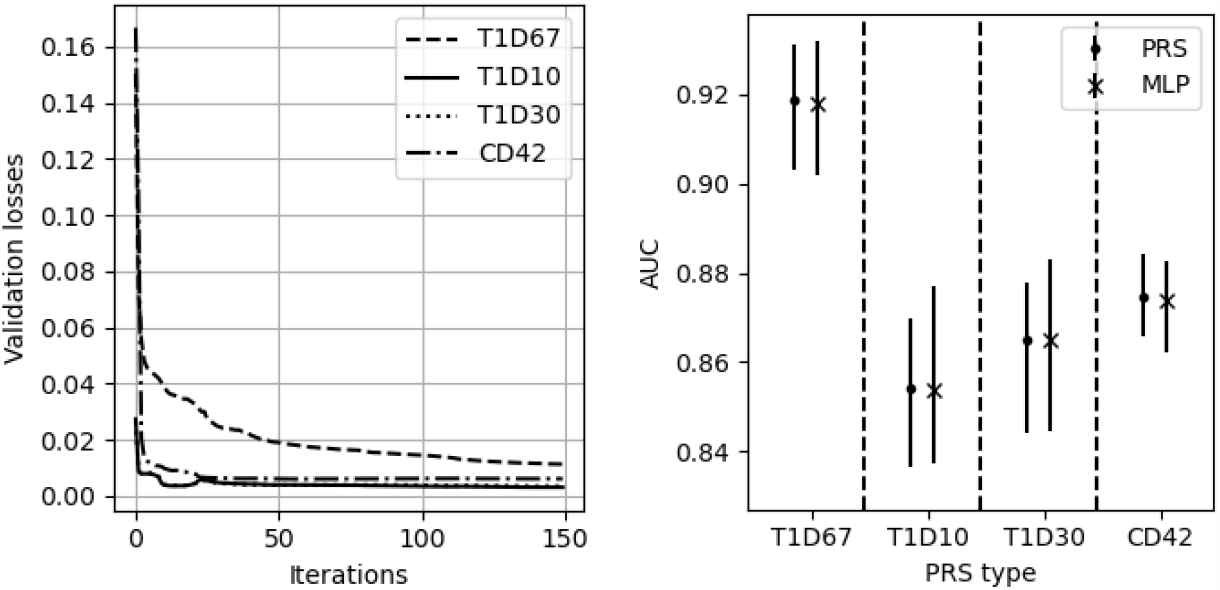
The performance of the multi-predictor MLP. Left) The validation losses (mean squared errors) of the multi-predictor MLP for each of the predicted PRSs. Right) The AUCs with uncertainties for the original PRSs and the multi-predictor MLP (denoted as MLP).

**Figure 8.**
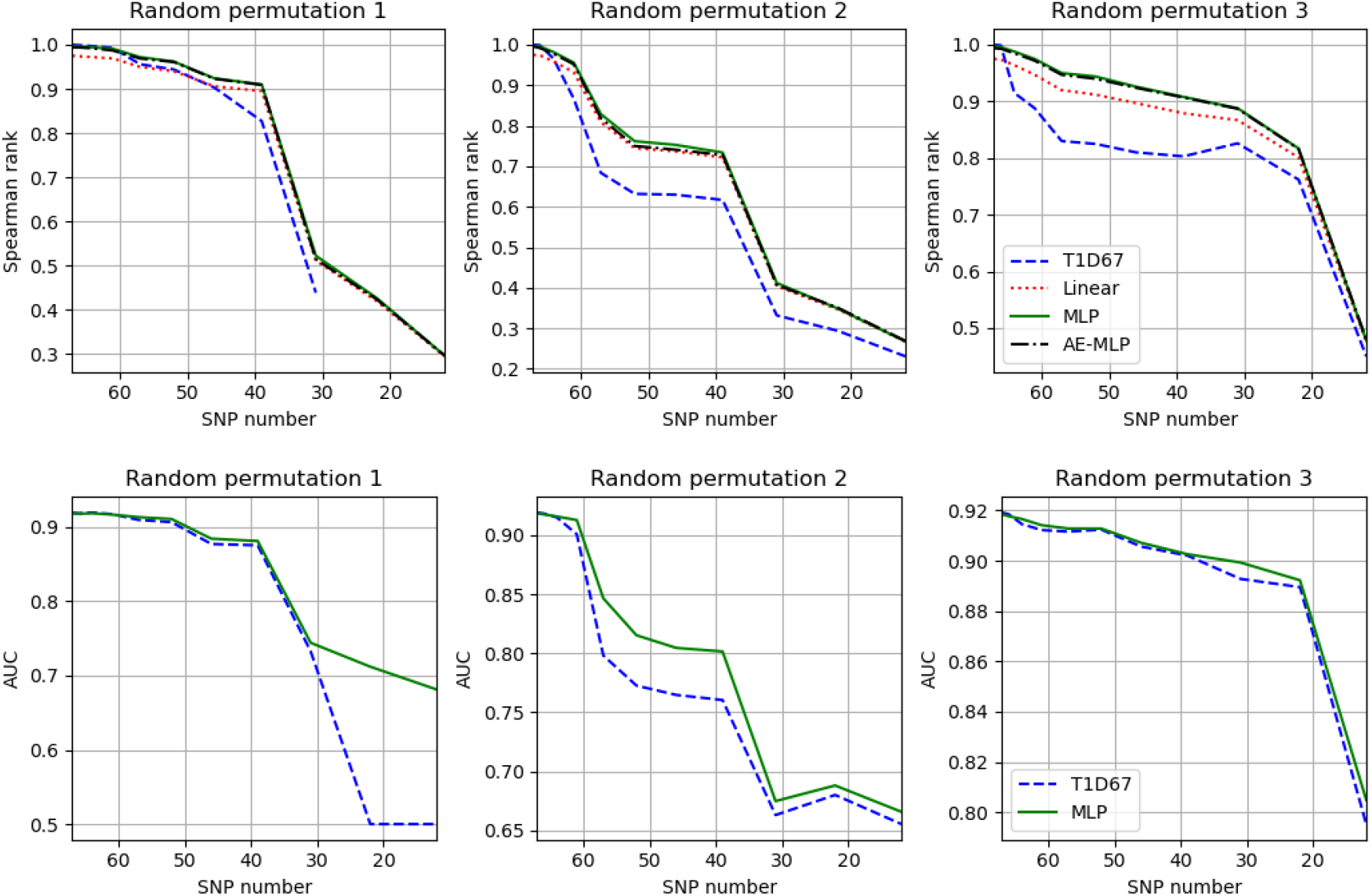
Top) Change in Spearman rank correlation coefficient for the T1D67, linear predictor, MLP and AE-MLP models with reducing number of SNPs. The three plots show different random permutations of removal of the SNPs. Bottom) The change in AUC for the removal of SNPs for the T1D67 and MLP. The y-axis varies significantly for different permutations.

### 3.2 Estimating risk scores with missing input SNPs

Figure 8 we show how the Spearman rank correlation coefficient changes with reduced numbers of SNPs for the T1D67 PRS, linear, AE-MLP and MLP models for different random permutations of removal of SNPs. The MLP and AE-MLP produce similar results for all SNP subsets. The linear predictions perform slightly worse than the MLP and AE-MLP when the number of SNPs being removed is not too large and tend to converge as the remaining number of SNPs is small. The MLP either performs comparably or better than the PRS. For different choices of removal of SNPs there is considerable variation in performance between the PRS and MLP. We also show the AUC between cases and population test sets for the PRS and MLP in the bottom plots.

In Figure 9 we show one example of the potential use of the MLP approach for one subset of 57 SNPs. On the left plot we show the distributions of the risk scores for the PRS and the MLP (both with 57 input SNPs). The left violin plot of each pair shows the non-T1D population test set and the right shows the T1D test set. The middle plot shows the PRS generated with 57 SNPs against the PRS67 with red dots showing non-T1D samples from the population test set (a random sample of 387 for viewing clarity) and blue crosses the T1D samples. The right plot shows the same results but for the MLP. With 57 available input SNPs the PRS and MLP produce AUCs of 0.798 (95% CI 0.779-0.818) and 0.847 (95% CI 0.828-0.864) respectively.

**Figure 9.**
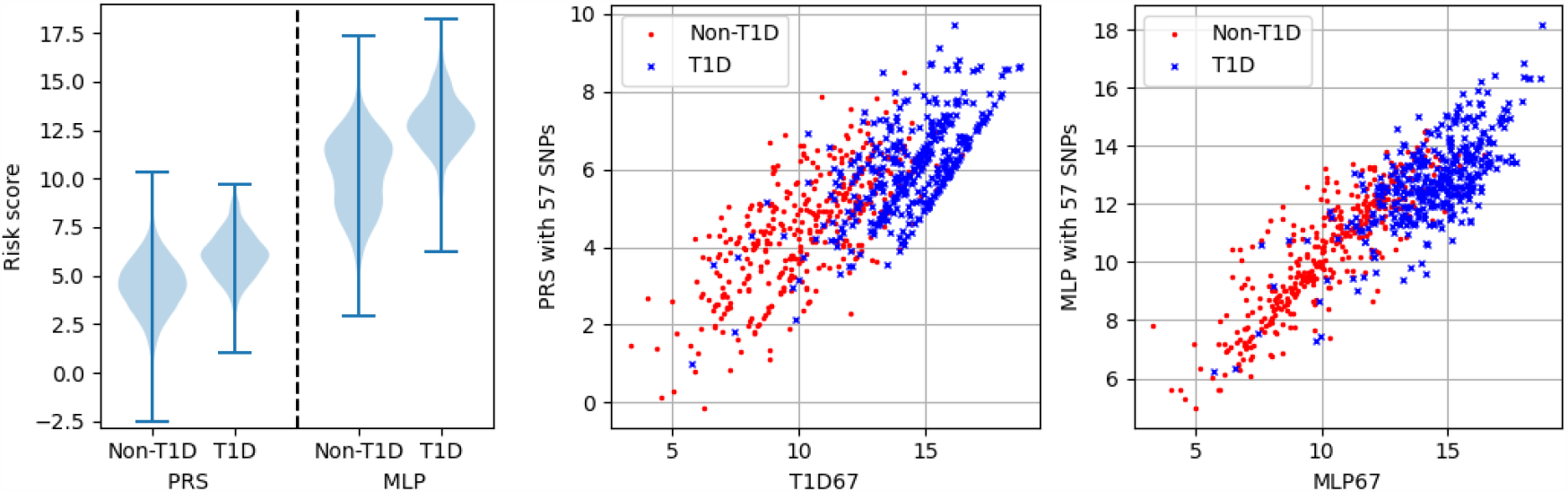
Example use case with 57 SNPs (10 removed). The AUCs for PRS and MLP on the reduced SNPs are 0.798 (95% CI 0.779-0.818) and 0.847 (95% CI 0.828-0.864) respectively. Left) Distributions of the non-T1D (left of the pair of plots) and T1D (right of the pair of plots) for the PRS (left pair) and the MLP (right pair). Middle) Plot of the PRS generated with 57 SNPs against the PRS with all 67 SNPs. Red dots are a random selection of 387 non-T1D scores and blue crosses the 387 T1D scores. Right) The same plots as the middle set but for the MLP.

The AE-MLP and MLP show very similar performance (see Figure 8) when input SNPs are unavailable. This suggests that the mechanism for the MLP is to estimate values (probabilities) of missing SNP values. We therefore trained the AE for all 67 SNPs with one held out each time, producing a prediction for each of the removed SNPs when all other 66 SNPs are available. We measure the performance for each removed SNP by the Pearson correlation coefficient and show a histogram of the results in the left plot of Figure 10. The most common result is that the AE cannot make any reasonable prediction of the SNPs (correlation below 0.1) but for some SNPs the AE performs well. The middle plot shows the AE performance on three types of SNPs in the 67 SNP PRS: those related to the HLA-DQ, HLA and non-HLA aspects of the genome. The AE performs well on the HLA-DQ SNPs, worse on the HLA SNPs and poorly on the non-HLA SNPs. As a demonstration of the capacity of the MLP to improve performance when the HLA-DQ SNPs are removed we train an MLP model removing a random permutation of those 14 SNPs. In the right plot of Figure 10 we show how the MLP performs compared to the PRS when these SNPs are removed.

**Figure 10.**
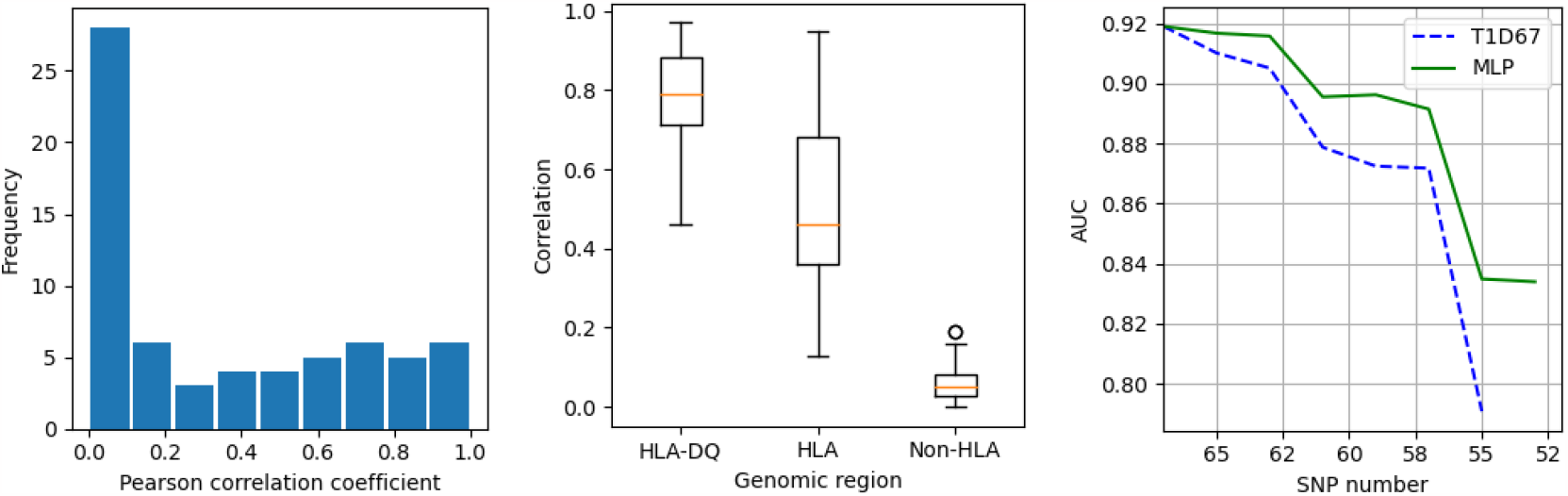
Left) A histogram showing the Pearson correlation coefficient between predicted and actual SNP values when we trained an AE with just the one SNP removed (67 models in total). Middle) The correlation of the AE predictions for SNPs in the HLA-DQ, HLA and non-HLA regions. Right) The AUC of the PRS and MLP against SNP number when the 14 SNPs in the HLA-DQ are removed in a random permutation.

We also show that this approach works when applied to the T1DGC dataset the model has not been trained on. In Figure 11 we show how the PRS and MLP performs for the reduction in SNP numbers as taken at random from the HLA-DQ set. In the left plot we show the AUC between the T1D and non-T1D partitions. The middle plot shows ROC curves for the results given by SNP number 57. The right plot shows violin plots of the risk scores produced by the PRS and MLP with 57 remaining SNPs.

**Figure 11.**
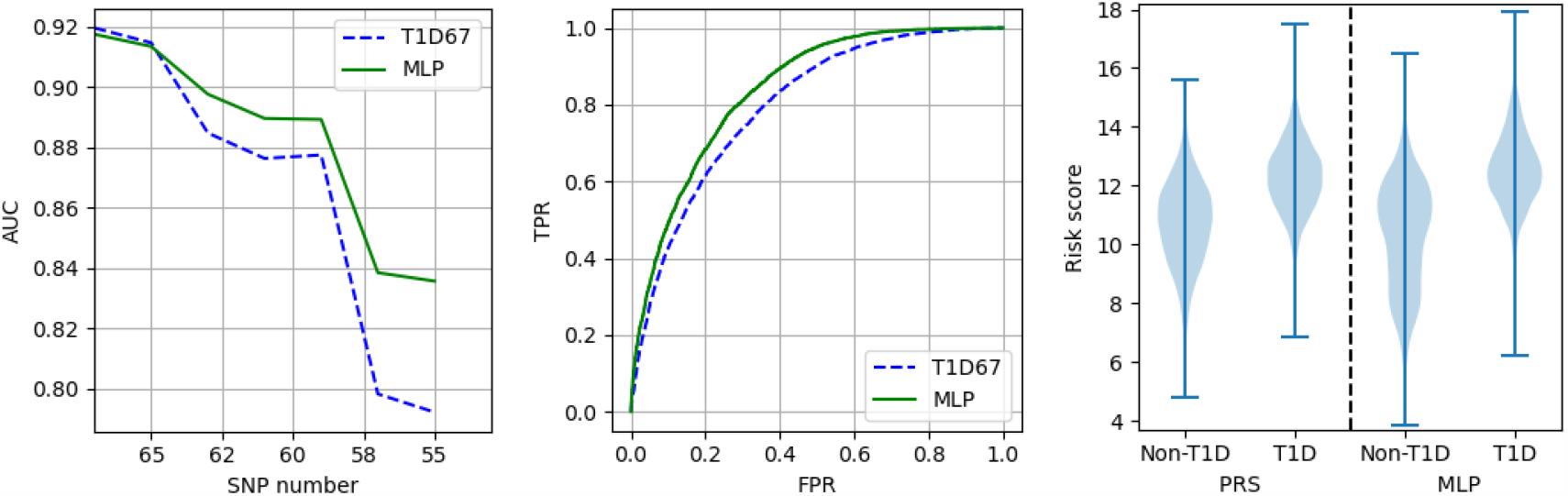
Results from applying models trained on the UKBB data to the T1DGC dataset. Left) AUCs between cases and controls when removing (at random) HLA-DQ SNPs. The vertical line is the SNP set results then shown in the middle and right plots. Middle) ROC curves of the MLP and PRS when 57 SNPs remain. Right) Distributions of the PRS and MLP scores for the controls and cases with 57 SNPs.

### 3.3) Exploring prediction quality limitations with missing SNPs

To investigate whether it is possible to improve on the baseline MLP performance with missing SNPs we consider one subset of 52 SNPs (the 52 SNP subset shown in the middle plot of Figure 8) and train the model variants discussed in Section 2.5. The metrics with uncertainties for these models are shown in Figure 12. All the additional models show similar performance with no statistically significant improvements over the standard MLP. In addition, a simple ensemble approach was performed taking the average of predictions from sets of all the models and no significant improvement in performance was observed.

**Figure 12.**
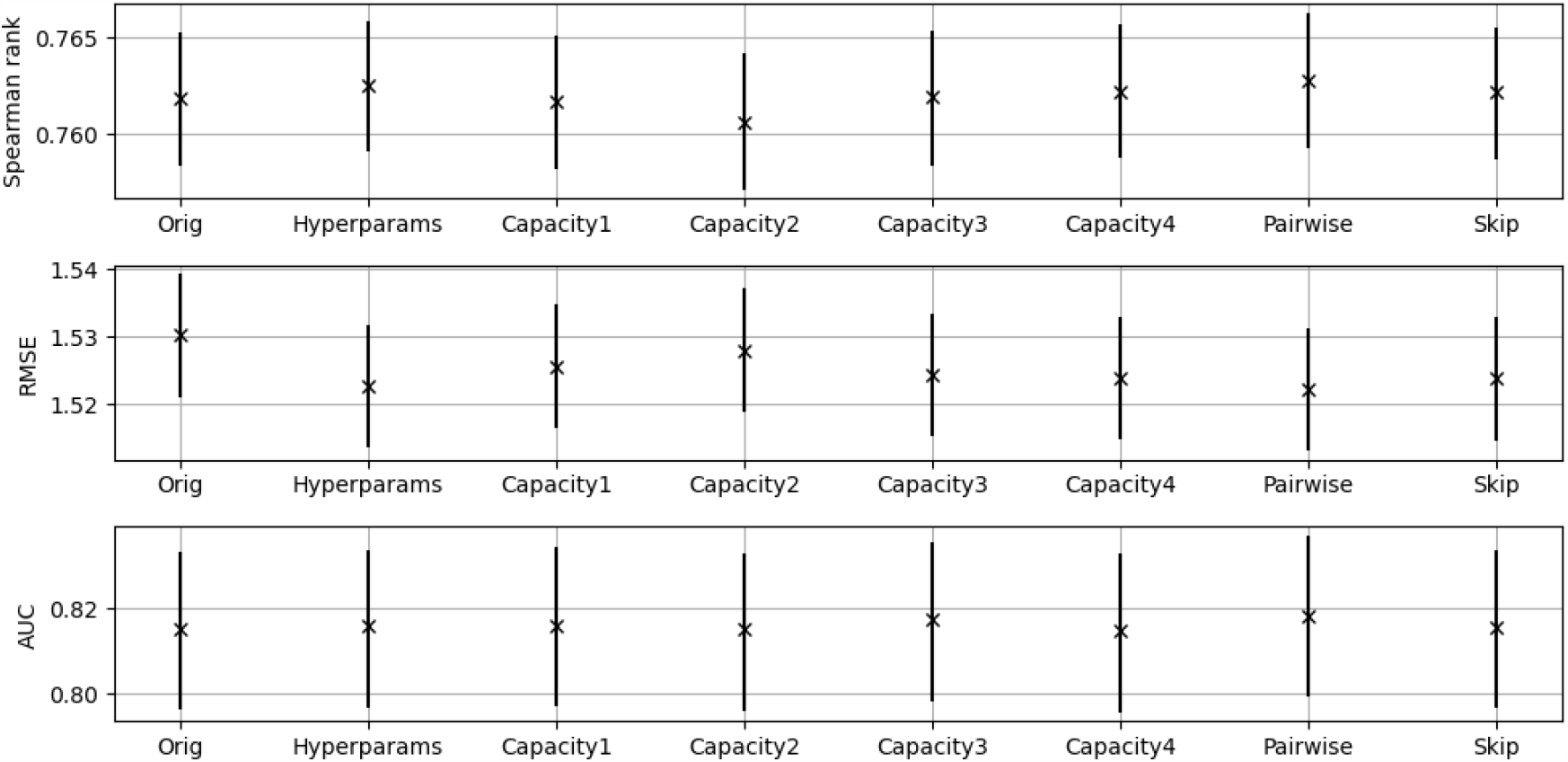
The performance of the additional models discussed in Section 2.5. The Orig denoted model is the standard MLP and the other models were named in Section 2.5. The Spearman rank correlation coefficient (Spearman rank) and RMSE are to the original PRS.

In Figure 13 we show predictions from a random 500 samples from the test set and plot the additional model predictions against the original model (Orig) predictions. The similarity between all additional models and the original predictions are high.

**Figure 13.**
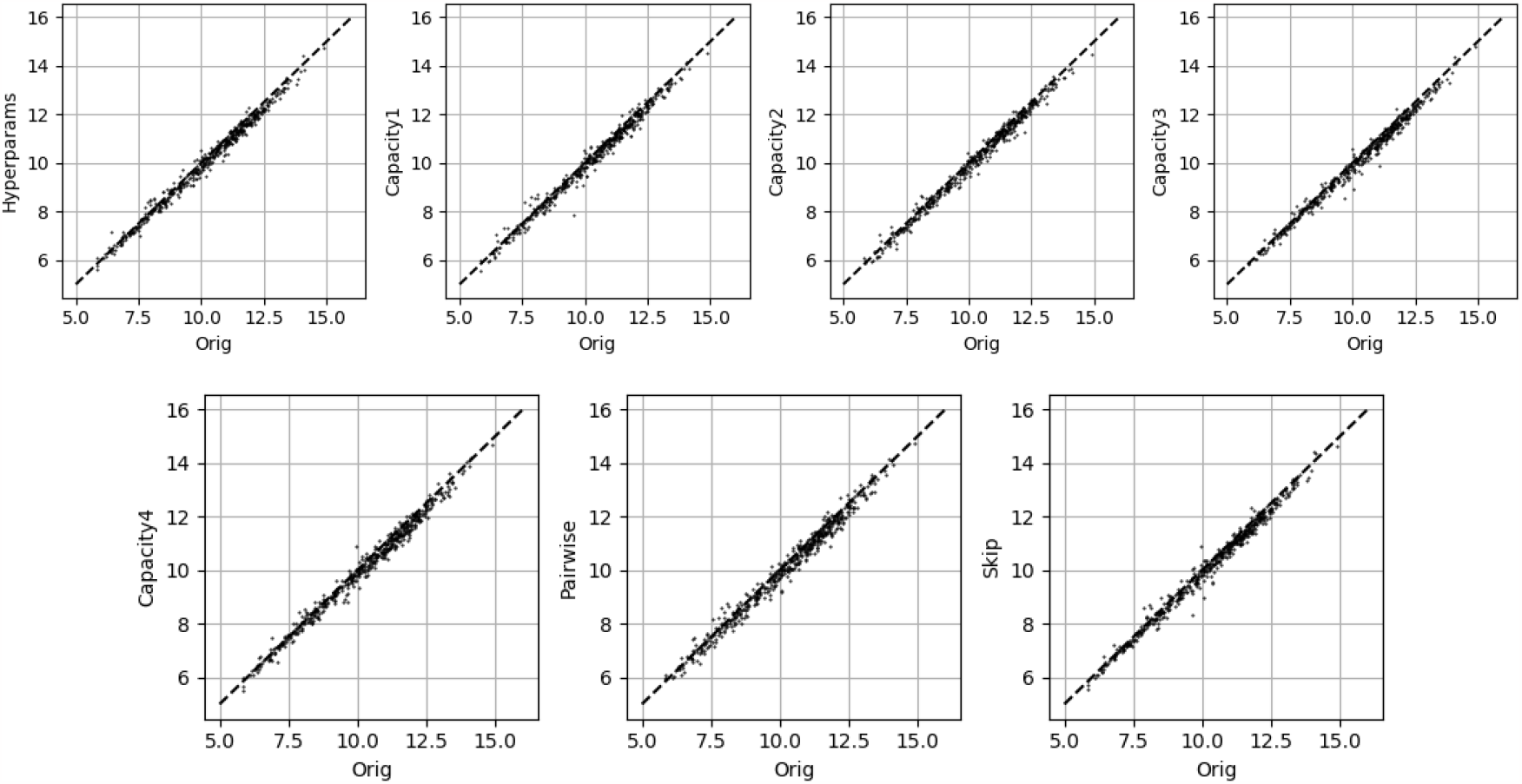
A random selection of 500 predictions on the test set for the additional models against the standard MLP (Orig). All models were trained on the same 52 input SNPs.

## 4) Discussion

We have shown that neural networks can be used to generate PRSs that match those of specifically written programmes when all input SNPs are available and that one NN-PRS can generate multiple PRSs with no loss of performance. One benefit of a neural network generated PRS (NN-PRS) is these can be easily transferred for use in other contexts. Instead of rewriting code the NN-PRS is a combination of matrix multiplications with a ReLU. To produce the four PRSs all that is required is the ordered list of input SNPs along with the weights. The PRSs can be generated in many different programming languages including using spreadsheets. This approach also has considerable longevity because the mapping from the input SNPs to the output PRS is via matrices which can be stored in simple formats such as a text file. There is therefore limited risk of changes in computing, programming languages, or updated technology which would make this form of model storage obsolete.

A second use of a NN-PRS is when considering the integration with other data to produce further predictive models. A standard PRS produces just one risk score output but the NN-PRS has a latent layer before the output neuron which encodes the input SNP information in a reduced dimensional vector rather than a scalar. For predictive models this latent encoding may prove more informative than just the summary scalar. Further models could use the latent vector as in input which, when combined with other data, could provide improved model performance.

Another use is in the burgeoning field of self-supervised learning (SSL). In SSL deep learning model are pretrained on pretext tasks so that when the model is finally trained on the desired downstream task it enhances its performance. A PRS has the potential to be a useful pretext task for a model to set the model weights effectively so the performance on the real task improves. This approach can either use the entire network or take parts of it as the pretrained parts for another model such as an improved risk generator.

PRSs are often produced when some of the input SNPs are unavailable. An neural network PRS (NN-PRS) has the potential to improve the quality of the risk score in these circumstances. We tested both an MLP and an AE-MLP on their capability to compensate for missing input SNPs, both performed at the same level. This similarity in performance suggests that the neural networks are predicting the scores of the missing SNPs (with some probability). Due to this similarity the performance of the AE at predicting missing SNPs shows when the NN-PRS will outperform the PRS with missing SNPs.

The AE can estimate some of the missing SNPs, with others not predictable. The NN-PRS therefore tends to operate at the equivalent level as the PRS (if it cannot estimate the missing SNPs) or outperforms the PRS. Most of the SNPs that are predictable tend to be related to the HLA-DQ and if SNPs are missing there then the MLP is likely to improve on performance.

An example use-case of the NN-PRS when used with missing SNPs was shown where the AUC of T1D to non-T1D was statistically significantly higher. In either research or clinical settings the difference in performances may be worth using the NN-PRS over a standard PRS.

We also demonstrated that these results are not due to overfitting to the specific training dataset. We applied the trained models to the T1DGC dataset, as an independent test set, and showed results that are very similar to those from UKBB.

One challenge with the missing SNP NN-PRS is that the potential combination of missing SNPs for a 67-SNP PRS is large and each SNP subset has to be trained separately. If there are a collection of SNPs that are likely to be missing then it would be possible to train a collection of models for those specific missing SNPs but it is not generalisable beyond that.

The alternative use-case is generation of the NN-PRS for each specific set of missing SNPs. This type of on-the-fly analysis is possible for two reasons. First the models are not difficult to train, we demonstrated that multiple different hyperparameter choices: learning rates, batchsizes and model architectures all produce similar final performance as long as the model architecture has a reasonable capacity. Consequently, a set of models could be set up to train automatically with the best automatically selected on a validation set. As the models are small and computationally inexpensive the multi-model approach is not costly. Secondly, we showed that the amount of training data needed is in the thousands which would only require a few MB of data storage. For a final use-case of this approach data could be bundled into the software package, models trained automatically on the data with known output PRS. The quality of the model could be directly provided to the user via the validation set and models produced for those specific available SNPs. Alternatively, anyone with access to a reasonable sized dataset could train these models specifically on their data for use on other datasets where the full set of SNPs is not available.

We also investigated the limitations of the NN-PRS to predict PRSs when SNPs are missing. We explored a range of variations to the MLPs trained on one specific SNP subset. We found no statistically significant improvement in performance via any of the approaches we took including the use of averages. Moreover, the final predictions made by all the models are very similar which suggests a convergence of model estimates. This similarity in performance provides evidence that the models are producing as good a set of predictions as are possible to make. As NNs are universal function approximators, and there are many more samples than input dimensions, this therefore suggests that there are no models that could further improve when the SNPs are missing. To further improve the prediction of the PRSs with missing SNPs likely requires addition inputs to the model rather than any alterations to the models.

## 5) Conclusions

MLPs can be easily trained to generate complex PRSs with equivalent performance to the coded models. A single MLP can produce multiple PRSs. These models are combinations of matrices with simple non-linearities that can be run on almost any analysis platform and require only simple storage. This approach should enable more researchers/clinicians to generate PRSs and effectively requires no maintenance of code.

When there are missing input SNPs the MLP can either match or improve on PRS performance. The level of improvement depends on the ability of the model to predict the values of the missing SNPs (with much higher performance for the HLA-DQ SNPs) combined with the importance of the SNP.

The models require modest amounts of training data to produce high performance and the training required does not need significant hyperparameter tuning. Therefore, these models on missing SNPs can be trained as they are needed depending on which specific SNPs are missing.

The evidence is that no further improvements in PRS estimation from missing SNPs is possible without the inclusion of additional information. Multiple variants of the models were trained all which produce similar performance and very similar specific predictions.

## Data Availability

All data produced are available online at https://www.ukbiobank.ac.uk/ and https://repository.niddk.nih.gov/studies/t1dgc/

## Acknowledgements

RAO and MNW had a UK Medical Research Council (MRC) Confidence in Concept grant to develop a type 1 diabetes GRS biochip with Randox and have ongoing research funding from Randox R&D. SS is funded by a grant from Randox.

SS, MNW and RAO are supported by the National Institute for Health and Care Research Exeter Biomedical Research Centre. The views expressed are those of the author(s) and not necessarily those of the NIHR or the Department of Health and Social Care.

RAO has research funding from a Diabetes UK Harry Keen Fellowship (16/0005529), National Institute of Diabetes and Digestive and Kidney Diseases grants (NIH R01 DK121843–01 and U01DK127382–01), JDRF grants (3-SRA-2019–827-S-B, 2-SRA-2022–1261-S-B, 2-SRA-2002–1259-S-B, 3-SRA-2022–1241-S-B, and 2-SRA-2022–1258-M-B) and the Larry M. and Leona B. Helmsley Charitable Trust.

## Code availability

Code is available at https://github.com/stevensquires/

